# What is the probability that this patient, who presents to a UK hospital, will be diagnosed with Covid-19? Prospective validation of the open-source CovidCalculatorUK resource

**DOI:** 10.1101/2020.12.06.20243691

**Authors:** George Chapman, Lewis Mundell, Charlotte H. Harrison, Tamsin Cargill, Odhran Keating, Mark Johnson, Andrew Smith

## Abstract

**Introduction:** The novel coronavirus SARS-CoV2 and the associated disease, Covid-19, continue to pose a global health threat. The CovidCalculatorUK is an open-source online tool (covidcalculatoruk.org) that estimates the probability that an individual patient, who presents to a UK hospital, will later test positive for SARS-CoV2. The objective is to aid cohorting decisions and minimise nosocomial transmission of SARS-CoV2.

**Methods:** This n = 500 prospective, observational, multicentre, validation study compared the CovidCalculatorUK’s estimated probability of Covid-19 with the first SARS-CoV2 oropharyngeal/nasopharyngeal swab result for individual patients admitted to hospital during the study period (01.04.20 − 18.05.20). A comparison with senior clinicians’ estimates of the probability of Covid-19 was also made.

**Results:** Patients who were prospectively grouped, by the CovidCalculatorUK, into 0-30% estimated probability, 30-60% and 60-100% estimated probability went on to have first swab SARS-CoV2 positive results in: 15.7%, 30.5% and 61.9% of cases, respectively. CovidCalculatorUK performance demonstrated an area under the curve of 0.76 (95% CI 0.71 – 0.81) (p < 0.001). Senior clinician stratification of the estimated probability of Covid-19 performed similarly to the CovidCalculatorUK.

**Conclusion:** The CovidCalculatorUK provides a reasonably accurate estimate of the probability of an individual testing positive on their first SARS-CoV2 nasopharyngeal/oropharyngeal swab. The CovidCalculatorUK output performs similarly to a senior clinician’s estimate. Further evolution of the calculator may improve performance.

## Introduction

The novel coronavirus SARS-CoV2 and its associated disease, Covid-19, continue to pose a global health threat. Throughout 2020, UK hospitals have treated exceptional numbers of patients with Covid-19^1^. Given the infectious nature of Covid-19, patients with suspected or confirmed Covid-19 are often admitted to predetermined wards, in a process known as “cohorting”. The Centres for Disease Control (CDC) define cohorting as: “the practice of grouping together patients who are colonised or infected with the same organism to confine their care to one area and prevent contact with other patients”^2^. In UK hospitals, nosocomial SARS-CoV2 transmission occurred in large numbers during the first wave before specific control measures were introduced^3^.

RT-PCR (reverse-transcriptase polymerase chain reaction) testing on nasopharyngeal/oropharyngeal swabs, or sputum/deep tracheal samples, is the current gold standard diagnostic test^4-6^. During the first wave of Covid-19 in the UK the delay from taking a swab in hospital to receiving the result ranged from one to three days^7^, though more rapid options are being explored^8^. During this time, whilst cohorting was being used to minimise nosocomial transmission, the uncertainty regarding the possible diagnosis of Covid-19 led to difficult clinical decision-making regarding the optimal ward for individual patients that required admission to hospital^9^.

The “CovidCalculatorUK” is a simple, open-source, not-for-profit online tool (https://CovidCalculatorUK.org/) that provides an estimate of the probability that an individual patient will later test positive for SARS-CoV2. The calculator is based upon published international data on SARS-CoV2 diagnostic features, including cough, fever, radiological findings and results from the full blood count. The calculator input is seven simple questions, the output is an immediately-available estimation of the probability (expressed as a percentage) that an individual patient will later test positive for SARS-CoV2. The CovidCalculatorUK was launched on 31^st^ March 2020, in the early phase of the first wave of Covid-19 in the UK.

Many diagnostic aids for triaging suspected Covid-19 cases are available^10^, including programs/apps^11-13^, online resources^12,14^, scoring systems^15-17^, decision trees^18^ and nomograms^19^. Jehi et al. 2020 describe an open-source online calculator with a similar output to the CovidCalculatorUK, though with a focus on community prediction of Covid-19 diagnoses^14^. Though many are available, the authors are not aware of any diagnostic aid using the method employed in the present study, which is based upon the JAMA Rational Clinical Examination Bayesian methodology^20^. Of the available diagnostic aids, few offer the simplicity and rapidity of the format used by the CovidCalculatorUK – allowing results to be gained in under 15 seconds of data entry from any online device. To the knowledge of the authors, this is the first study that includes and compares estimates produced by a Covid-19 diagnostic tool, estimates produced by senior clinicians, SARS-CoV2 swab results and the cohorted location of the patients.

We present an examination of whether the CovidCalculatorUK produces an accurate prediction of the first SARS-CoV2 swab result, as well as a comparison with senior clinicians’ judgements. The wider objective is to aid and inform cohorting decisions and minimise nosocomial transmission of SARS-CoV2.

## Methods

The method used to create the CovidCalculatorUK is described in supplementary file 1. This prospective, observational, validation study gathered data from three NHS hospitals within the UK, two in England, one in Scotland. At site one 38 days between 01.04.20 and 18.05.20 were selected (chosen for research team availability) and all patients admitted through the medical “take” on the selected days were included in the analysis. At site two and three all medical patients admitted in the defined period were included (site two 23.4.20 − 28.4.20, site three 20.3.20 - 20.04.20). The availability of the research team defined the study period at each site. The validation study ran until 500 patients were included in the analysis. Temperature (assessed with an infra-red tympanic thermometer) and fraction of inhaled oxygen were recorded from the first set of observations taken upon arrival to hospital. Presence or absence of cough was assessed by the clerking doctor. Whether the chest radiograph or computed tomography scan of the chest were deemed “abnormal” was recorded according to the judgement of the clinical team.

Within eight hours of admission, the research team used the CovidCalculatorUK to estimate the probability of Covid-19 for each individual and recorded this data. Senior clinicians (Consultants, Registrars & SAS Doctors) did not have access to this data at the time. The senior clinicians were asked about their feelings regarding the probability of Covid-19 (in two formats: binary yes/no and in graded probability: no suspicion/some suspicion/high suspicion) in the individual in question and the data recorded. If the clinical team was unavailable the medical notes were examined to ascertain the documented suspicion of Covid-19 and the cohorting location. Clinicians’ assessments and cohorting locations were available at study site one and two only.

Patients were excluded from the validation analysis if there was insufficient clinical data. As a minimum, for the data entry to the calculator, the presence/absence of fever and cough had to be recorded. Patients were excluded if there was a known positive SARS-CoV2 swab in the 14 days prior to admission. Hospital protocol, at all three sites (during the study period), dictated that SARS-CoV2 swabs be sent for all medical admissions. SARS-CoV2 RT-PCR swab results were reviewed 10 days later, and the results of any swabs within that time period entered into the dataset. Patients without a swab result during the 10-day period were excluded from the study.

Statistical analysis (SPSS V26, Excel 2016) was performed by grouping the CovidCalculatorUK output into estimated probability groups of 0-30%, 30-60% and 60-100% (group thresholds were determined post hoc). Division into three categories was chosen to mirror clinical protocols categorising patients into low, medium and high risk, or by the red-amber-green “traffic light” system. The analysis assessed accuracy of prediction when compared with the observed test results using the current gold standard test (RT-PCR of nasopharyngeal/oropharyngeal swabs), as well as comparison with clinician estimates. Laboratory assay used: Altona RealStar SARS-CoV-2 Assay. Primary analysis focused on the first SARS-CoV2 swab for each included patient. Receiver operating characteristic (ROC) curves were used to explore the optimal cut-offs (of the calculator output) to categorise individuals/groups most effectively. Student’s t-test was calculated for difference of means, Mann-Whitney U test for ROC AUC analysis and χ^2^ for 2×2 tables.

## Results

The CovidCalculatorUK estimated the probability of Covid-19 in 529 individual medical patients admitted to three hospitals in the UK (246 patients from site one, 44 from site two and 210 from site three). The predefined exclusion criteria resulted in the following exclusions: five patients with positive SARS-CoV2 swab results prior to admission, four patients with insufficient clinical notes and 20 patients without a SARS-CoV2 swab sent to the laboratory. Following these exclusions, 500 patients remained in the completed dataset.

Of the 500 patients, 141 had a positive first SARS-CoV2 swab result, giving a prevalence in this validation cohort of 28.2%. A further 7 patients (with a negative first SARS-CoV2 swab) had a positive result on their second SARS-CoV2 swab (using the definition of any positive SARS-CoV2 swab in the first 10 days of admission gives a study Covid-19 prevalence of 29.6%).

**Table 1.** **Comparison of patient groups with first SARS-CoV2 swab positive and first SARS-CoV2 swab negative:**

|  | Positive first SARS-CoV2 swab | Negative first SARS-CoV2 swab | All patients |
| --- | --- | --- | --- |
| Admission temperature >37.5°C | 82/135 (60.7%) | 96/321 (29.9%) | 178/456 (39.0%)* |
| Cough | 102/135 (75.6%) | 157/321 (48.9%) | 259/456 (56.8%)* |
| Abnormal CXR | 108/132 (81.8%) | 134/288 (46.5%) | 242/420 (57.6%)* |
| Abnormal CT chest, within 48 hours of admission | 7/8 (87.5%) | 21/49 (42.9%) | 28/57 (49.1%)* |
| Required supplemental O2 (% of patients)<br>(mean FiO2) | 30/47 (63.8%)<br>(FiO2 = 0.36) | 57/199 (28.6%)<br>(FiO2 = 0.26) | 87/246 (35.4%)<sup>†</sup><br>(FiO2 = 0.28) |
| Mean white cell count x10<sup>9</sup>/L (IQR) | 9.9 (6.5 - 15.0) | 10.3 (7.1 - 12.8) | 10.2 (6.9 - 13.0)<sup>†</sup> |
| Mean lymphocyte count x10<sup>9</sup>/L (IQR) | 1.3 (0.6 - 1.4) | 1.4 (0.6 - 1.6) | 1.4 (0.6 - 1.6)<sup>†</sup> |
| CovidCalculator output, estimated probability of Covid-19 as a percentage. Group mean shown. | 52.2% (IQR 12.8% - 88.0%)<br>95% CI for the mean (46.2% - 58.1%)<sup>‡</sup> | 20.6% (IQR 2.0% - 37.5%)<sup>‡</sup><br>95% CI for the mean (17.8% - 23.3%) | 29.5% (IQR 2.5% - 54.4%)** |

\* data from site one and site three
<sup>†</sup> data from site 1 only
<sup>‡</sup> p < 0.001 (t-test)
\*\* all 3 sites included, n = 500 datapoints

**Table 2.**
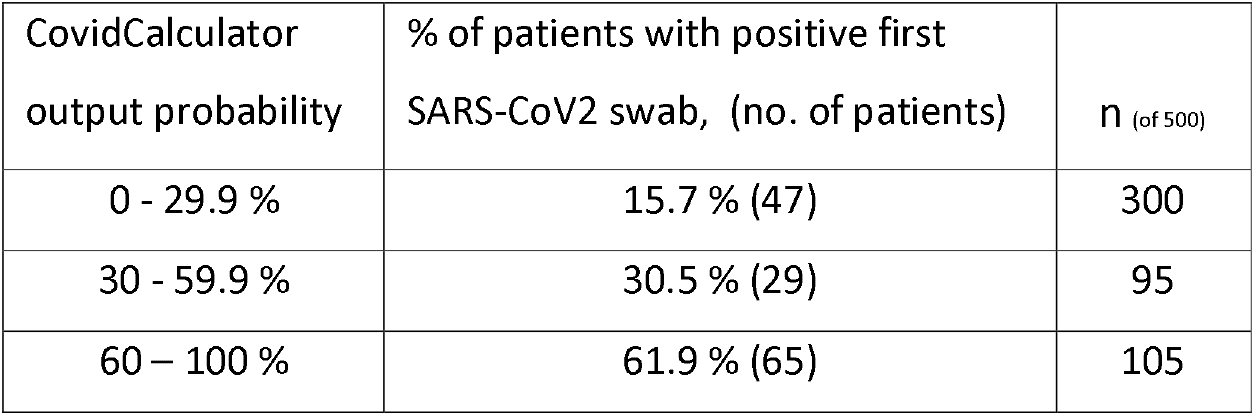
**Calculator output (estimated probability of Covid-19 as a percentage) compared with first SARS-CoV2 swab result:**

**Fig 1.**
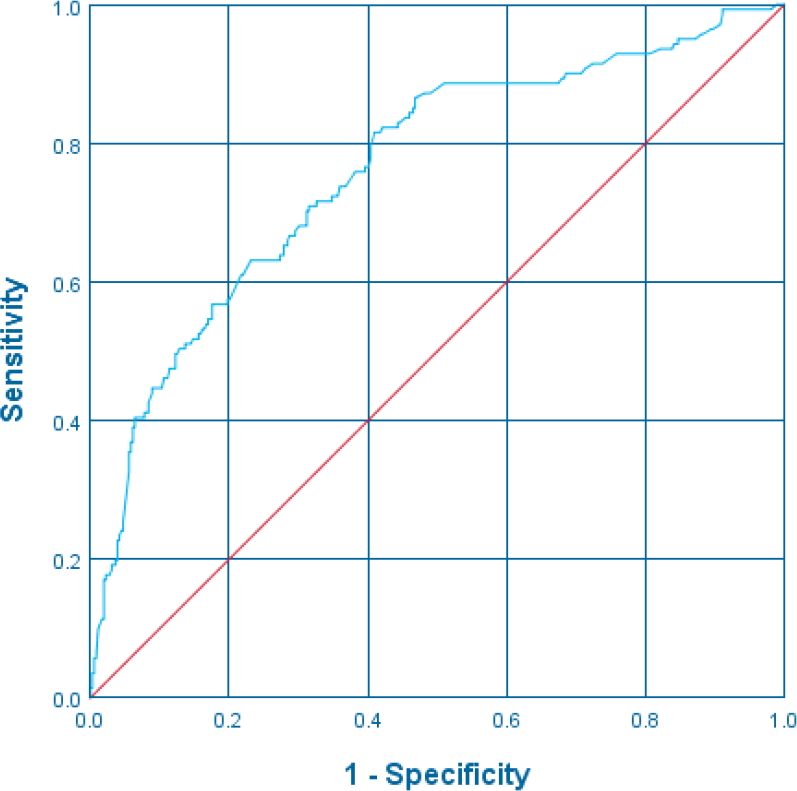
Receiver operating curve for the CovidCalculatorUK output shows general performance of the model, (for predicting first SARS-CoV2 swab result) with an area under the curve of 0.76 (95% CI 0.71 – 0.81) (p < 0.001). Further expression of the performance and output of the calculator can be found in supplementary file 2.

**Table 3.** **Demonstrating CovidCalculator cut-offs to predict first SARS-CoV2 swab result:**

| CovidCalculator cut-off defining group | n (of 500) | Within the defined group, % of patients with positive first SARS-CoV2 swab, (no. of patients) | Sens. | Spec. | PPV | NPV |
| --- | --- | --- | --- | --- | --- | --- |
| > 5% | 322 | 38.8% (125) | 0.89 | 0.45 | 0.39 | 0.91 |
| > 15% | 228 | 44.7% (102) | 0.72 | 0.65 | 0.45 | 0.86 |
| > 30% | 200 | 47.0% (94) | 0.67 | 0.71 | 0.47 | 0.84 |
| > 45% | 149 | 53.7% (80) | 0.57 | 0.81 | 0.54 | 0.83 |
| > 60% | 105 | 61.9 % (65) | 0.46 | 0.89 | 0.62 | 0.81 |
| > 75% | 77 | 71.4 % (55) | 0.39 | 0.94 | 0.71 | 0.80 |
| > 90% | 48 | 68.8% (33) | 0.23 | 0.96 | 0.69 | 0.76 |

**Table 4.** **Senior clinicians’ stratifying patients into three categories of estimated probability of Covid-19*:**

| Clinician classification<br>defining group | n<br>(of 290) | Within the defined group, % of<br>patients with positive first SARS-CoV2 swab, (no. of patients) | Sens. | Spec. | PPV | NPV |
| --- | --- | --- | --- | --- | --- | --- |
| a. "No clinical suspicion" | 114 | 5.3 % (6) | 0.46<sup>†</sup> | 0.89 | 0.95 | 0.27 |
| b. "Some clinical suspicion" | 98 | 15.3 % (15) | 0.28<sup>‡</sup> | 0.65 | 0.15 | 0.80 |
| c. "High clinical suspicion" | 78 | 41.0 % (32) | 0.60<sup>‡</sup> | 0.81 | 0.41 | 0.90 |
| "Some suspicion" or "high<br>suspicion" (b and c) | 176 | 26.7 % (47) | 0.89<sup>‡</sup> | 0.46 | 0.27 | 0.95 |

\* Available for site one and site two.
<sup>†</sup> For the "no clinical suspicion" category, the sensitivity, specificity etc. are for detecting a negative first SARS-CoV2 swab result.
<sup>‡</sup> For the "some clinical suspicion and "high clinical suspicion" category, the sensitivity, specificity etc are for detecting a positive first SARS-CoV2 swab result.

**Table 5.**
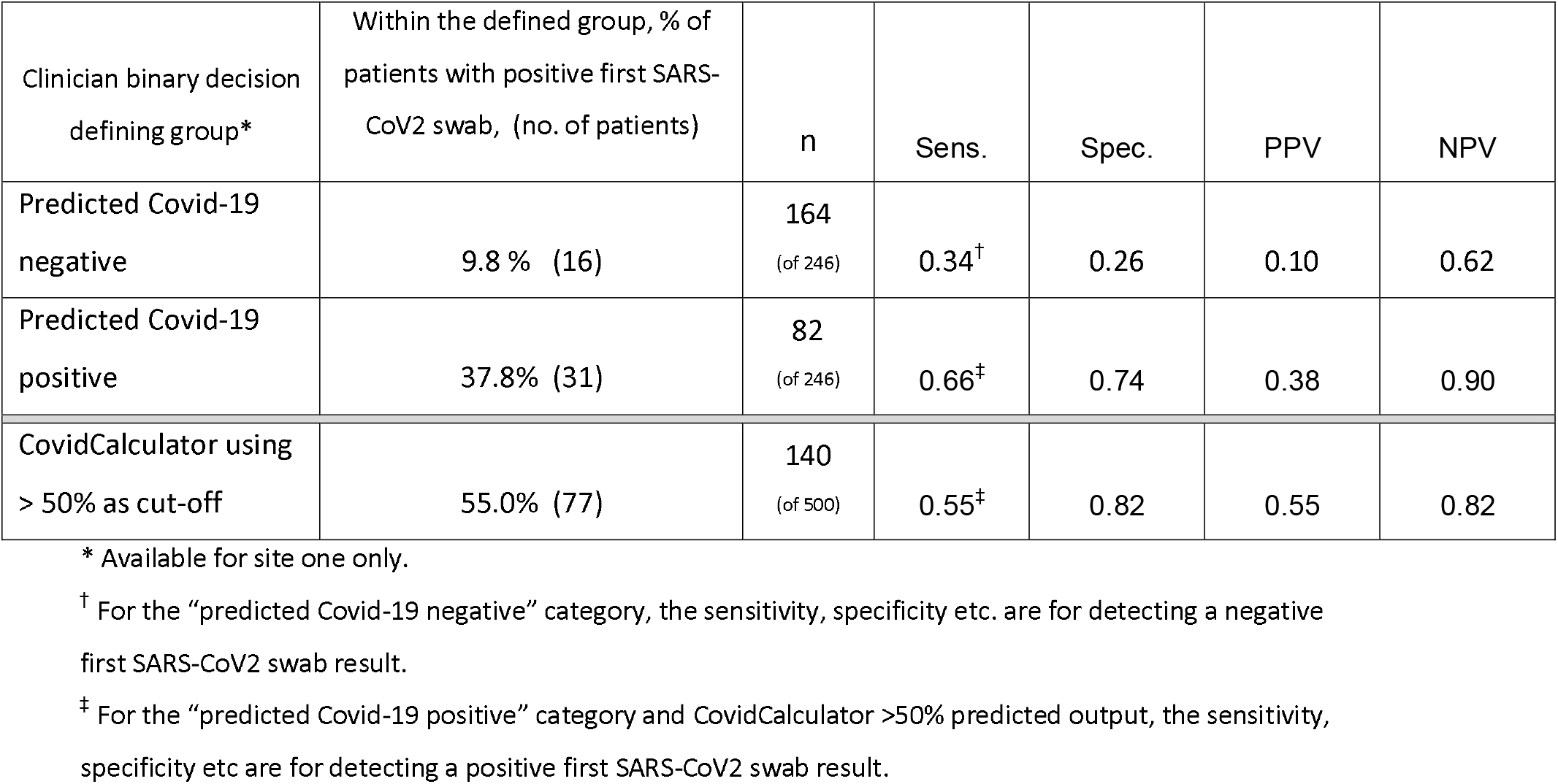
**Senior clinicians/CovidCalculator outputs, for binary decisions:**

Though the majority of clinicians’ predictions were made by Consultants (220 of 246 predictions, data regarding seniority of clinician available for site one only), the remaining predictions were made either by Registrars or SAS doctors. If the clinician predictions are classified as “correct” (i.e. if a positive SARS-CoV2 swab result was predicted and this later proved true, or a negative SARS-CoV2 result was predicted and this later proved true), then the proportion of “correct” predictions over the total number of predictions can give an idea of how well the patients were categorised by their SARS-CoV2 status. Using this measure, Registrars/SAS doctors made “correct” predictions 69.2% of the time, compared with Consultants, who made “correct” predictions 73.2% of the time - this slightly improved performance was not statistically significant (p = 0.67 by χ^2^) when comparing the number of correct and incorrect predictions between Consultants and Registrars/SAS doctors.

Using this same measure, if the CovidCalculator cut-off value of 50% estimated probability of Covid-19 was used to categorise patients as predicted positive or negative, then this prediction was “correct” 74.6% of the time. If the cut-off value of 75% was used, then the prediction was “correct” 78.4% of the time. There was no statistically significant difference in performance for either of these cut-offs, by this measure, when compared with senior clinicians (p > 0.1 by χ^2^).

### Cohorting patients

The data collected at site one and two recorded the clinicians’ verbal or documented feelings regarding the probability of Covid-19 in individual patients – this was gathered solely for the purpose of this study. The actions clinicians took in deciding which “cohort” to place a patient in was also recorded. There were 289 cohorting actions made at site one and two. 144 patients were cohorted to Covid-19 areas (i.e. “red” or “amber” areas), 145 were cohorted to non-Covid-19 areas (i.e. a “green” area). Of the 145 cohorted to a non-Covid-19 area, there were 12 positive first SARS-CoV2 swabs (8.3% of patients). Of the 144 cohorted to a Covid-19 area, there were 41 positive first SARS-CoV2 swabs (28.5% of patients). 174 of 289 (60.2%) cohorting decisions were, by this definition, “correct”. This is a highly simplified model of cohorting and not a balanced representation of the true clinical picture, which utilised side rooms, barriers within bays and more graded risk assessments within clinical areas.

### Alternative definition for Covid-19 cases - any positive SARS-CoV2 swab in the first 10 days of admission

Of the 500 included patients, there were seven cases where the first SARS-CoV2 swab result was negative and a further swab result in the following 10 days was SARS-CoV2 positive. Re-analysis of the data presented in the current study with this altered definition had no clinically significant effect upon the results.

## Discussion

Predicting the diagnosis of Covid-19 in patients presenting to hospital has challenged clinicians throughout 2020. This prediction, and the cohorting of patients to reduce the transmission of SARS-CoV2 in hospitals, is vital. The CovidCalculatorUK resource provides an estimated probability that an individual patient, assessed on admission, will go on to test positive for SARS-CoV2. The current study sought to prospectively validate the output of the CovidCalculatorUK resource.

The results of this validation study show that the CovidCalculatorUK estimate of the probability of Covid-19 in the group that went on to have a first SARS-CoV2 test positive was significantly different to the group that went on to test negative (mean estimated probability 52.2% vs 20.6% respectively, p < 0.001). Using a CovidCalculatorUK threshold of greater than 90% estimated probability detected patients that would later test positive for SARS-CoV2 with a specificity of 0.96 (PPV 0.69). Using a threshold of greater than 5% estimated probability detected patients that would later test positive for SARS-CoV2 with a sensitivity of 0.89 (NPV 0.91). The AUC across the range of estimated probabilities showed reasonable performance, at 0.76 (95% CI 0.71 – 0.81). If binary predictions are classified as “correct” if a positive SARS-CoV2 swab result was predicted and this later proved true, or a negative SARS-CoV2 result was predicted and this later proved true, then Consultants made “correct” decisions 73.2% of the time. The CovidCalculatorUK (at a cut-off of 50% estimated probability) binary prediction was “correct” 74.6% of the time.

An important added consideration, with Covid-19 diagnostic predictions, is the proportion of asymptomatic individuals with SARS-CoV2 infection, estimated to vary between 5% and 80%^21^. Presence or absence of cough and fever are important inputs into the CovidCalculatorUK - as a result there is a risk of underestimating the probability of Covid-19 in asymptomatic patients. Indeed, the sensitivity of the CovidCalculatorUK for the detection of patients that would later test positive for SARS-CoV2 was lower than expected. Perhaps this is a reflection of the performance of the calculator itself, or due to the proportion of patients with an asymptomatic or atypical presentation of Covid-19. Added to the challenge of asymptomatic presentations is the false negative rate from swabs tested using the RT-PCR method – this was estimated to be 20-40% during the first wave of Covid-19 (though this does appear to vary greatly depending upon the day of SARS-CoV2 exposure and the day the SARS-CoV2 swab was collected in the individual in question)^22^. The current study shows that the “expected” outcomes estimated by the CovidCalculatorUK were generally higher than the observed outcomes from swab results (as groups: Estimated probability 0-29.9%: 15.7% positive observed; 30-59.9%: 30.5% positive; 60-100%: 61.9% positive). This result may be explained by the CovidCalculatorUK producing overestimates, and/or a tendency for SARS-CoV2 RT-PCR swab tests to under-report true positives.

The current study is not unique, sitting within a sphere of similar prediction tools. However, the use of the Bayesian method in Covid-19 prediction is believed to be, at the time of writing, unique. The use of an estimate of regional population prevalence (albeit regional hospital presenter prevalence) as the starting point for further calculations means this method is well suited to adapt to the dynamic nature of a pandemic - where population prevalence in a given region can vary by an order of magnitude in a matter of weeks. This method does have its shortcomings: it is only as accurate as the data available to it – much of which is, in itself, estimated. The likelihood ratios for the symptoms and investigations are based upon published data for a population outside of the UK – the assumption that this data can be directly applied to the UK population may have compromised accuracy. Though the current study has focused on the diagnosis of Covid-19 in UK hospitals, the method described here could potentially be applied to any region with a known hospital presenter Covid-19 prevalence and has potential utility in a range of international settings.

Many tools that aid in predicting the diagnosis of Covid-19 have been made available in 2020, using a variety of methods and user interfaces. The resource with the greatest similarity to the current study was produced by Jehi et al. and gives an online open-source estimate, based on a calibrated nomogram approach from 18 possible data entry inputs^14^. The AUC from the validation cohort in Jehi et. al was 0.84. This study focused more heavily on screening and testing in lower probability groups in the community. Increasing in complexity the Soltan et al.^13^ artificial intelligence modelling, using electronic patient records, vital signs, blood gas results and laboratory values (for patients admitted to hospital, incidentally in the same region of the UK as site one and two of the current study) yielded an impressive AUC of 0.87 in its validation cohort, a PPV of 0.4 and NPV of 0.98. Many other diagnostic tests have focused exclusively on radiological findings^10^. The CovidCalculatorUK does integrate chest radiographic assessment and CT chest assessment – interestingly the former was almost ubiquitous, whilst the latter was rarely included in the calculations on admission – perhaps due to the time delay often associated with acquiring computed tomography scans during the Covid-19 pandemic. Simplicity and rapidly-available estimates are a central feature of the CovidCalculatorUK – in competition for simplicity of prediction with the current study is the elegant PARIS score. Tordjman et al. assign zero, one or two points based on cut-off values of the eosinophil, lymphocyte, neutrophil and basophil counts to assign patients as low, intermediate or high probability of Covid-19. This method achieved an AUC of 0.89 in their validation cohort – though the prevalence of Covid-19 in the retrospective validation cohort was 69%, which may elevate the sensitivity and specificity found during validation. The authors of the current study believe the use of regional prevalence as the starting point for further calculation in the CovidCalculatorUK generates more reliable results across a range of population prevalences, as compared to the nomogram/algorithm-based approaches - where the sensitivity and specificity may vary dramatically depending upon the population the tool is applied to^13^.

There has been an abundance of new diagnostic or prognostic aids during the Covid-19 pandemic^10^. Set against the need for rapid innovation in the face of a new clinical problem is the need for sound statistical analysis and the appropriately cautious use of novel unvalidated tools. Though the current study does describe a novel prediction tool, the method of creating the tool is fully described (Supplementary file 1) and the reasonably large prospective validation in 500 patients in three centres seeks to provide sufficient evidence for an external judgement to be made on the utility of this novel tool.

When compared with other published prediction tools, the CovidCalculatorUK does appear to be less accurate. Balanced against this diminished accuracy is the simplicity, relative paucity of data required, easy availability and rapidity with which the CovidCalculatorUK can be used. The format of the output, as an estimated probability expressed as a percentage, has utility for application to cohorting policy that is flexible to the hospital space/s available and disease prevalence in a local area. The current study has been purposefully silent on clinical recommendations for cohorting or setting thresholds where certain actions are recommended - as this will depend upon the number of cohorting options and facilities available at each site, as well as population factors. The statistically optimal cohorting strategy is clearly complex. Blanket recommendations are unlikely to have universal clinical utility. Best practice in this area is centred around the “enhanced traffic care bundle” – based upon the successful measures used in the SARS outbreak of 2003^23,24^. This traffic care bundle is a more practical organisational approach, with less emphasis on the individual decisions regarding patient allocation to different zones of the hospital. A core difficulty is in balancing the sensitivity and specificity of a predictive or diagnostic test for Covid-19. A highly sensitive, less specific test risks the Covid-19 cohort “red” zone containing SARS-CoV2 negative patients (and therefore highly susceptible to nosocomial infection), with a relatively safer “green” zone. By contrast, a highly specific, less sensitive test ensures the “red” zone contains predominantly Covid-19 patients, though the “green” zone, as a result of the compromised sensitivity (in favour of specificity) is likely to house SARS-CoV2 positive infectious individuals placing a greater number of susceptible patients at risk of nosocomial infection. Though the “susceptible-infectious” model is well established^25,26^ - the optimal statistical strategy to balance these risks and minimise nosocomial transmission in various hospital spaces is sorely needed during the current Covid-19 pandemic.

The method used by the CovidCalculatorUK draws on the well-established Bayesian methodology described by the JAMA rational clinical examination series^20^, building a post-test probability from the prevalence in the hospital-presenter population and known symptoms/investigations of the disease in question. This method iterates towards a diagnosis through information sequentially added into consideration. Though Bayesian theory is not overly welcomed in clinical medicine, many believe that clinicians naturally formulate their diagnoses, using their clinical experiences, using this same iterative method and Bayesian inference^27,28^, even if it is not labelled as such. The current study demonstrates, for the first time, that it is possible to apply this method, with reasonably accurate results, to predict the diagnosis of a novel disease in individual patients. This study also demonstrates the performance of clinicians undertaking the same task. It is interesting to note the marked similarities between the CovidCalculatorUK and clinician estimates, though these estimates were generated entirely independently of one another. Their similarity may allude to the common ground shared between a calculated Bayesian method and clinical decision-making amongst senior clinicians.

Perhaps the similarity in performance, when compared with senior clinicians, means the CovidCalculatorUK tool has limited additional clinical utility. In the presence of senior clinicians this may be true, though the calculator can produce rapidly equivalent results in the absence of senior clinicians, on a large scale, throughout the day and night or in remote settings – where senior clinician input may not be readily available. Anecdotally, from both Consultant colleagues and other healthcare professionals, the calculator has provided a welcome second assessment. It is noteworthy that in the early phase of the present study, clinicians were asked to estimate the numerical probability of Covid-19 in individual patients – many had difficulty with this request, favouring verbal descriptions of probability, such as “very unlikely”. Subjective statements such as these have been shown to vary greatly in meaning between individuals^29^.

Prediction of SARS-CoV2 infection status has wide-reaching impact. Though the current study has focused on the probability of testing positive and the influence upon cohorting - there are other practical concerns - including whether relatives/contacts should be advised to self-isolate and operational considerations, such as oxygen utilisation and hospital/staff resourcing.

The authors hope the need for this work, and the CovidCalculatorUK resource, will diminish as accurate near-patient tests become available, with possible results in 30-90 minutes^8,30^. If these tests perform perfectly, the uncertainty in cohorting will be eliminated and nosocomial transmission dramatically reduced. However, if concerns remain regarding false negative results in patients felt to have a high pre-test probability, the uncertainty in cohorting will remain. In such scenarios, the pre-test probability remains necessary to place the result of the test in context and facilitate further decision-making. The CovidCalculatorUK can calculate the influence of a negative or positive SARS-CoV2 swab (LR 0.41 and LR 47.5 respectively) on the estimated probability of Covid-19. This feature was not validated in the current study due to the circularity of incorporating swab results into the calculation. The SARS-CoV2 RT-PCR swab results, as the current gold standard test, were required as the independent comparison with the CovidCalculatorUK output.

## Conclusion

The present study describes a method for estimating the probability, using readily available parameters, of an individual patient testing positive for SARS-CoV2 following admission to hospital. Successfully cohorting patients during the first wave of Covid-19 in the UK has been challenging. The goal of the present study was to validate the output of the CovidCalculatorUK resource and in doing so, prepare for further work seeking to mathematically optimise cohorting strategy and minimise the transmission of SARS-CoV2 within hospitals. The open-source CovidCalculatorUK resource (covidcalculatoruk.org) performed similarly to senior clinicians’ estimates, both producing reasonably accurate predictions of which individuals would later test positive for SARS-CoV2. Further evolution of the calculator will aim to improve performance.

## Data Availability

Data referred to in this manuscript is not currently available in the public domain. This is in keeping with the ethics approval granted for this study.

## Acknowledgements

The authors would like to thank the Academic Health Sciences Network, including Mamta Bajre and Guy Checketts for their assistance in the development of this study. We also wish to thank Raha West, Chris Cleaver and William Clackett for their assistance during this study. Many thanks to the Mercedes-AMG Petronas Formula 1 team, including Serge Daval, Andrea Semprebon, Thomas Sutton and Michael Taylor for their assistance with the potential development beyond this initial project to future versions. Thank you to Andrew Chapman for his guidance and assistance in the statistical analysis. And a final heartfelt thank you to all of our colleagues in the NHS.

## Abbreviations

Covid-19: Coronavirus disease - 2019
SARS-CoV2: Severe acute respiratory syndrome coronavirus 2
RT-PCR: Reverse transcriptase polymerase chain reaction
CI: Confidence interval
CDC: Centres for Disease Control and prevention
JAMA: Journal of the American Medical Journal
RCE: Rational Clinical Examination
NHS: National Health Service
UK: United Kingdom
SAS: Specialty and associate specialist doctors
SPSS: Statistical package for the social sciences
IQR: Interquartile range
ROC: Receiver operating characteristic
AUC: Area under the curve
Sens: Sensitivity Spec: Specificity
PPV: Positive predicted value
NPV: Negative predictive value
LR: Likelihood ratio
FiO2: Fraction of inhaled oxygen
CXR: Chest radiography
CT: Computed tomography

