## Supplementary File 1 for "What is the probability that this patient, who presents to a UK hospital, will be diagnosed with Covid-19? Prospective validation of the open-source CovidCalculatorUK resource"

**Creating the CovidCalculatorUK**

The calculator uses the method described by Prof. Sackett in the JAMA rational clinical examination series^20^. The Bayesian method makes use of the formula: post-test odds = pre-test odds x likelihood ratio.

An approximation of the daily prevalence of Covid-19 in the accident and emergency (A&E) presenter population, on any given day or group of days, is required. This data was sourced from the Gov.UK Covid-19 “dashboard”^1^. The mean number of daily positive tests in the preceding three to seven day period (three days was used in the first phase of the first wave - as the cases were rising sharply – when case rates fell a weekly approach was used) served as the numerator. This data was from hospital laboratory testing, and was later called “pillar 1” when other forms of testing were introduced. This numerator was divided, for each region of the UK, by the number of A&E attendances per day in recent weeks (from Gov.UK). The availability of up-to-date total A&E attendances varied by country. Attendances in England were from available published data from the preceding month, Scottish and Welsh A&E attendances from the most recently published three weeks and N. Ireland from the same time period in 2019. From this data the pre-test odds in a given region could be crudely estimated from the total number of Covid-19 diagnoses made from hospital labs divided by the total number of A&E attendances.

The described core symptoms of Covid-19, including cough and fever, as well as radiological findings, findings from the full blood count and biochemical data were all investigated for their ability to discriminate between Covid-19 and other causes of presentation to A&E.

Following exploration of the published data at the time, the following inputs into the calculator were added.

The likelihood ratios, for each of these inputs, were calculated by comparing the historical attributes of Covid-19 negative UK A&E presenters (from published studies in prior to 2020) and comparing them to published presenting attributes in Covid-19 positive patients presenting to international hospitals^31^.

Fever: Reference values (4.4-8.5% adults presenting to A&E)^32-34^, Covid-19 positive values (43.8% on admission)^31^.

Cough: Reference values (1.4-3.9% in adults presenting, though as cough is rarely the primary symptom it is poorly coded, 8-10% A&E presentations in 2019 with acute respiratory infections)^35^. Covid-19 positive values^31,36^.

Chest radiograph abnormality: Reference values^37^, Covid-19 positive values^31^.

Lymphocyte count: Reference values^38,39^, Covid-19 positive values^31^. (50-60% of general medical patient admitted present with lymphopaenia).

White cell count: Reference values^38,40^, Covid-19 positive values^31,41^.

The decision was taken to use WCC for the less unwell and lymphopaenia for the more unwell given the finding that more severe presentations could demonstrate higher WCC^41^, aiming to avoid underestimating the risk in more unwell patients. WCC and lymphopaenia are not independent tests given one is formed in part from the other, as a result, the CovidCalculatorUK separated into two streams, more unwell (as judged by fraction of inhaled oxygen) and less unwell, the former using lymphopaenia and the latter using WCC.

Computed Tomography of the chest: values^42,43^.

RT-PCR: This featured as the final input of the CovidCalculatorUK. If a known swab was available the probability could be adjusted in line with this result^42-45^. During the validation of this study this was not analysed as the goal was in prediction for patients with an unknown status. This was included to emphasise the importance of a positive result in influencing the probability of a patient being infected with SARS-CoV2, but also the relatively lesser importance of a negative swab result in the pre-test probability was very high (given the relatively high false negative rate seen during the first wave^46^).

Anosmia featured more heavily as a core symptom of Covid-19 later in the first UK wave of the pandemic. Accurate data for the prevalence of anosmia in the A&E presenter population was not available, and as a result the likelihood ratio would likely have been inaccurate and anosmia did not therefore feature. The feeling of breathlessness was explored but was not felt to be of benefit to the calculator. Similarly, lactate dehydrogenase (LDH) levels are often high in patients admitted to hospital and exploration of its inclusion yielded poor differentiation of SARS-CoV2 positive and SARS-CoV2 negative patients. Consideration was made for the inclusion of gastrointestinal symptoms into the calculator, however their performance in discrimination between SARS-CoV2 positive and SARS-CoV2 negative patients was not sufficient for inclusion. Despite the absence from the calculator, physicians would be wise to consider “atypical” gastrointestinal presentations of Covid-19 in their patients, in order to prevent transmission of Covid-19 between patients e.g. on surgical wards not expecting Covid-19 patients^47^.

2 x 2 tables for each CovidCalculatorUK input, as well as sensitivity, specificity and likelihood ratio if present and if absent provided below.

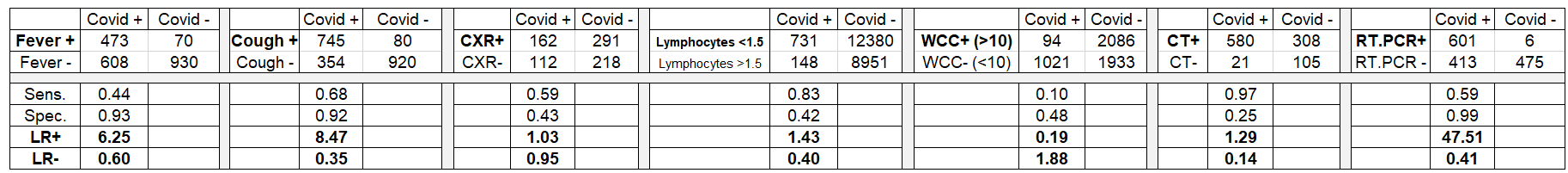
Table 1.

The resulting inputs for the online calculator included: date of the patient’s presentation, region in the UK, temperature on admission ( >37.5^o^C), cough, abnormal chest radiograph, inhaled oxygen (>50% or <50%), white cell count and lymphocyte count. Whether the CT scan of the chest was abnormal and whether there was an existing SARS-CoV2 RT-PCR result could also be entered. Options for each clinical input included: “positive”/“negative”/“not done” for each input.

All inputs, with the exception on the date of presentation and region were optional, and an automatically updating output probability was provided to the user as the calculator’s inputs were added.

Practicalities of data entry:

Given the heterogeneity of Covid-19 appearances on chest radiographs, and the desire to have clear binary decisions when entering data into the CovidCalculatorUK – the decision was taken to use the broad and undefined term of “CXR abnormal”: Yes/No. This allows the user to exercise their judgement, rather than attempting to clearly define the multitude of changes associated with Covid-19. A similar approach was taken regarding CT chest results. The finding of non-Covid-19 abnormalities such as lung malignancies can confound this system, though most users in the validation study described these cases as CT “normal” – choosing to read the Calculator’s query as “normal” indicating the absence of changes associated with Covid-19 and ignoring bystander pathologies.

Correcting for time lag to results:

Due to the delay between taking a swab to gaining the result and reporting the result to Gov.UK it was felt necessary to project the model forwards to compensate for this time lag. During the first wave of Covid-19 in the UK, the CovidCalculatorUK projected the A&E presenter population Covid-19 prevalence forwards by 72 hours, in order give a more accurate representation to clinicians entering data for patients admitted on the current day in question.

The first wave of Covid-19 in the UK was treated, for the purposes of this time lag calculation, as having three phases. Phase one, with a rising number of cases (31.3.20 – 10.4.20), phase two with cases plateauing (10.4.20 – 20.4.20) and phase three (20.4.20 – 5.7.20) with cases falling. These three phases were tracked from published Italian data (REF) and a best-fit relationship was generated. During phase one the chosen fit used a third order polynomial generated from the Italian data. During phase two and three, simple linear models were used. This polynomial/linear projection was applied to the most recently published UK cases data to extrapolate 72 hours ahead – in effect estimating the number of Covid-19 cases that would be present 72 hours after the publishing of the Gov.UK data. This projected number was divided by the currently published number of UK cases to give a simple “multiplier”. For simplicity, this multiplier was chosen to be a number, to two decimal places. Using this multiplier allows the testing lag to be “corrected” – giving an improved estimate of the current day’s Covid-19 prevalence in the A&E presenter population by accepting that the most recently published Gov.UK data is, at the time it is published, already approximately 72 hours out of date. Throughout the existence of the CovidCalculatorUK, this multiplier ranged between 0.95 and 1.2 inclusive. 1.2 used from 31.3.20 to 14.4.20, 1.0 - 1.15 used from 15.4.20 to 26.4.20 and 0.95 - 0.97 used from 27.4.20 to 5.7.20. This multiplier was updated daily. This multiplier is the only transformation or correction any of the data underwent in the CovidCalculatorUK (during the first wave).

This resulting combined calculator was translated into a user-friendly online interface, with easy to use toggle switches, accessible via mobile devices, tablet computers or desktop computers with access to the internet. (Fig. 1)

Fig. 1

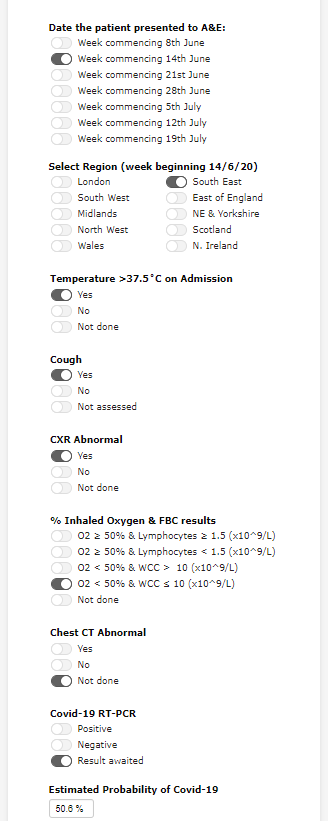

Using the method described, the CovidCalculatorUK estimates, for the current day/group of days, the A&E presenter prevalence of Covid-19 (the pre-test probability, which is then converted to odds) and by multiplying by serial likelihood ratios the calculator generates the posterior odds, for the individual in question, and converts this back to a probability, expressed as a percentage chance of that individual going on to test positive for Covid-19.

The online CovidCalculatorUK was launched, as an unvalidated tool, on 31.3.20.
