## Supplementary File 2 for "What is the probability that this patient, who presents to a UK hospital, will be diagnosed with Covid-19? Prospective validation of the open-source CovidCalculatorUK resource"

Histograms and BoxPlot expressing distribution of CovidCalculatorUK output for patients with negative SARS-CoV2 swab “0” and those with first SARS-CoV2 swab positive “1”.

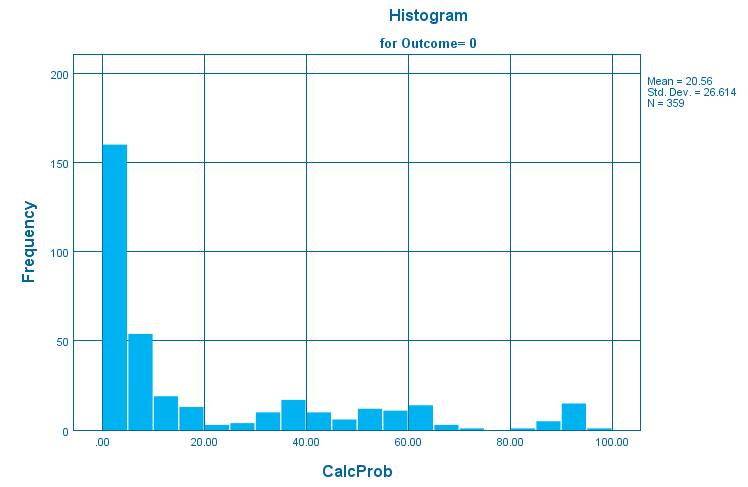

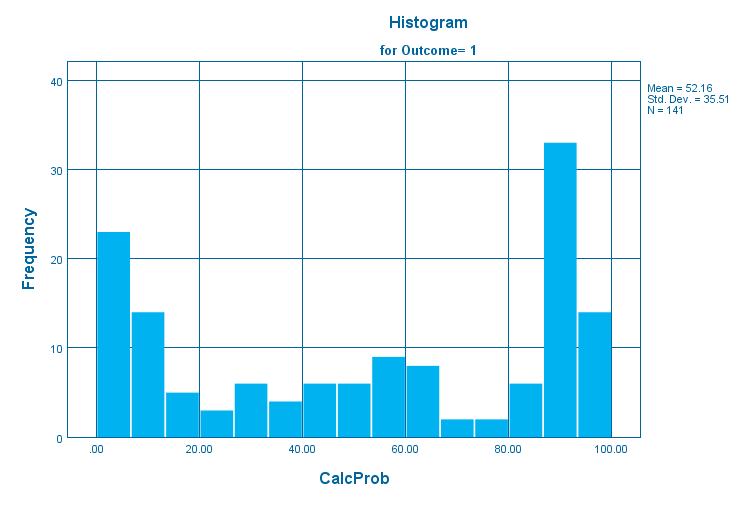

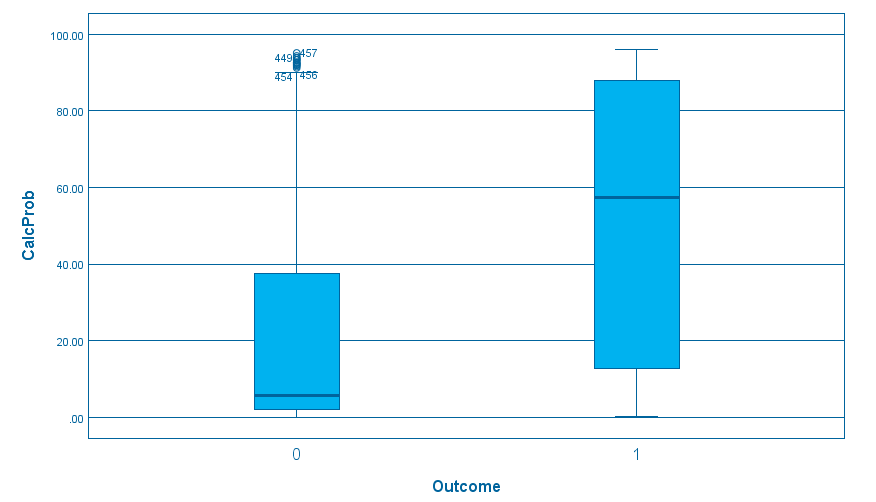
